# Evaluation of a brief intervention to reduce cell phone use in college students

**DOI:** 10.1101/19009241

**Authors:** Brian J. Piper, Shay M. Daily, Sarah L. Martin, Maurice W. Martin

**Affiliations:** Department of Medical Education, Geisinger Commonwealth School of Medicine, Scranton, Pennsylvania, United States of America; Department of Community Health Education, University of Maine at Farmington, Maine, United States of America; University of Southern Maine, Portland, Maine, United States of America; School of Pharmacy, Husson University, Bangor, Maine, United States of America

**Keywords:** adolescent, behavior, cognition, mobile phone

## Abstract

**Background:** Excessive cell phone use contributes to distracted driving, may increase risk for automobile accidents, and a minority of mobile phone users exhibit behaviors consistent with technological addiction. The purpose of this study was to determine whether cell phone beliefs and behaviors could be changed by a brief educational encounter. The Theory of Reasoned Action provided a lens for viewing attitudes and behavior.

**Methods:** A one-week pre-post design with a thirty-day follow-up was used with participants (N = 215, 67.0% female, age = 20.0 ± 1.6) assigned to a peer led intervention or comparison groups. The intervention included cell-phone educational materials. A short index of cell phone behavior was developed which showed good internal consistency with a Cronbach’s alpha of .81.

**Results:** The intervention group “agreed” or “strongly-agreed” more than the comparison group on five of the seven areas of cell phone beliefs and behaviors (*p* < 0.05, item Cohen’s *d* = .32 to .47, total *d* = .50) at one-week following receipt of informational materials.

**Discussion:** We conclude that attitudes and behaviors regarding cell phones are malleable and susceptible to change in young-adults following a brief psychoeducational intervention.

## Introduction

Most citizens of developed countries have access to and use cellphones, or similar mobile devices, to transfer information and data [1–3]. Cultural demands of the information age require most users to engage in use of mobile technologies to be a part of employment, education, recreation, or social-life [4–6]. Additionally, there are socially implied sanctions imposed for people who do not keep up with and use mobile technology frequently [7]. Mobile phones have replaced some other devices (e.g. cameras, address book) but may become an object of attachment [8].

There may also be adverse health consequences associated with cellphone use [9]. Glucose was elevated in the orbitofrontal cortex of adults on the side of the brain adjacent to a cell phone receiving a call [10]. These neural changes were temperature independent and glucose was reduced in the temporoparietal junction and anterior temporal lobe of young-adults (age 21 to 29) on the same side as cell phone exposure [11]. There is also an extensive body of data utilizing electroencephalographs [12]. A meta-analysis identified subtle, but significant, reductions in sperm motility (−8.1%) and viability (−9.1%) among mobile phone users [13]. One-hour of exposure to a mobile phone to young-adults in their early twenties increased plasma lipid peroxidases and decreased antioxidant enzymes [14]. Understanding whether these acute effects increase the susceptibility to certain diseases has been the topic of intense interest [15]. Mobile phone use does not appear to increase the risk for glioblastoma [16] and microarray studies have generally been unable to identify a proteomic profile that is beyond what would be expected by chance [17].

There are several epidemiological findings in this area which include, arguably, the best established and most consequential risks. There were 3,157 documented automobile fatalities which involved distracted driving in the U.S. in 2016 and, of these, a cell-phone that was in-use at the time of the accident for over four-hundred [18]. Even these numbers may be underestimates as they do not take into account how texting at night contributes to drowsy driving [19] which could further increase motor vehicle accidents. Two-thirds of adults (age 18-65) in the U.S. reported talking on their cell phones and one-third received or sent a text or email while driving in the past month [20]. Even for the minority who do not choose to use mobile phones, proximity to the devices carried by their peers and others can impose risks. Non-users can be the victims of injuries sustained by user inattention [21]. A video game for mobile devices has resulted in accidents [22]. On the other hand, public health investigations have generally found that text-messaging interventions can produce positive outcomes for diabetes self-management, weight loss, smoking cessation, and medication adherence [23,24].

There is a behavioral component to wireless technology use that may increase unintentional injury and mental health issues [25]. Overuse or misuse of this technology may have parallels to addiction [2, 26, 27] although much further research may be needed to establish this [28]. People with addictive behaviors are not always able to make rational decisions regarding that behavior [2, 29, 30]. For some users, an inordinate amount time is spent in non-productive activities that cut into time spent on healthier endeavors [21, 31].

The millennial generation, born between 1982 and 2002 [32, 33], may have a special relationship with technology [7, 22]. This is the first generation to grow-up with mobile information-based technology being likened to adequate educational preparation and success as an adult [34]. Educational leaders have spent an enormous amount of time, money, and administrative resources continuously upgrading modern education paradigms to procure technologically friendly environments [35, 36]. Millennials are more likely than prior generations to demand and use technology in innovative ways to enhance their lives [37, 38]. However, it turns out that their focus is primarily for social activities rather than academic or professional which is aided through their use of technology. Millennials, primarily traditional college age students, spent an average of 14 hours texting, 6.5 hours talking on the phone, and 6.5 hours using social media sites per week with some sending over 200 texts in a day [39].

The Theory of Planned Behavior has been refined by Ajzen and colleagues over the past four-decades [40–42]. Behavior is viewed as guided by beliefs about the consequences, the normative expectations of others, and the presence or absence of factors that may impact it. These behavioral beliefs, normative beliefs, and control beliefs contribute to the attitude toward the behavior, the subjective norm, and perceived behavioral control, respectively, which together result in behavioral intention [41]. The Ajzen model has been widely utilized in such diverse areas as nutrition [43], exercise [44], and responsible drinking [45].

It is currently unknown whether cell-phone attitudes and behavior are modifiable. Therefore, the purpose of this study was to explore the likelihood of behavior change regarding mobile devices following a brief educational intervention to millennials. This research question was embedded within the Theory of Reasoned Action and Planned Behavior [40–42].

## Materials and methods

### Participants

The participants were young-adult college students (N = 215, 67.0% female, age = 20.0 ± 1.6, Min = 17, Max = 28) at a public institution in northern New England.

### Procedure

Research team members held informal meetings to develop reliable quantitative surveys with items based in concepts from the Theory of Reasoned Action [40–42]. Drafts of each instrument were piloted using demographically similar non-participant groups.

A comprehensive search of the literature was conducted regarding the potential for negative health effects associated with cellular phone use including sleeplessness, injury and other situations and behaviors deleterious to human health. Using these results [6, 10–13, 20, 21, 25, 31, 46 – 50], a brief, one-page educational fact sheet was produced to use as an intervention tool. When instruments and tools were satisfactory to the research team, approval was obtained from a university IRB.

A quasi-experimental study design was employed. It was a one-week pre-post-intervention design with a subset of participants also completing a 30-day follow-up. Due to the potential benefits of the intervention, delayed intervention was used with the comparison group assuring the ethical treatment of all participants. The data collection protocol was determined by the research team and data collection packets were provided to each data collector. Due to the public-health (e.g. distracted driving) risks associated with cell-phone use, participants were assigned to the intervention or comparison groups in an approximately 3:1 ratio. Each data collection packet held a pre-coded participant log with pre-determined selected numbers identifying comparison group participants (participant #2, #5, #7, #14, #20, etc.), the informed consent, and 20 pre- and post-intervention coded surveys to provide identification for follow-up.

The one-page fact sheet was provided to the intervention group immediately after the pre-intervention survey was completed. The comparison group received it immediately after the second (i.e. one-month) post-intervention survey (See Appendix A for the instrument). There were seventeen volunteer data collectors who were trained to gain written informed consent, administer the survey, and provide the fact sheet effectively. Each data collector was charged to recruit twenty willing potential participants who would be assigned to either an intervention or a comparison group. All potential participants were “cold contacts” of the millennial generation (age < 30) attending a public liberal arts institution in the northeastern United States.

Recruitment commonly occurred in public areas. No incentives were offered for participation. As part of the pre-intervention survey, demographic and general cell phone information (cell phone type and basic usage frequency), were collected. The survey also included items based in the Theory of Reasoned Action and Planned Behavior, transitioning from values to attitude and intention such as: “I believe there may be negative health effects related to cell phone use” and “I should change some of my behaviors when it comes to my cell phone use” (Appendix A). One week later, the first post-intervention survey was administered to intervention and control groups. A subset of both groups completed the post-intervention survey at one-month after the baseline. The fact sheet was shared with the comparison group members at this juncture after the second post-test data had been obtained.

#### Statistical Analyses

Descriptive statistics were used to describe the characteristics of both groups. We compared cell phone use and attitudes and intentions for the two groups by calculating the percentage of agreement with survey items which was analyzed with a chi-square with Yates correction. Differences between groups on parametric measures were expressed in terms of Cohen’s *d*. Variability was expressed as the SD unless noted otherwise. Comparisons between the intervention versus the control group was analyzed in SYSTAT, version 13.1 (Chicago, IL) with alpha set at 0.05. Internal consistency was determined with Cronbach’s alpha. A total index of cell-phone attitudes and behaviors was calculated by summing the seven post-test ratings (1 = strongly agree to 5 = strongly disagree), with the two negatively worded items reverse scored. Figures were prepared with GraphPad Prism, version 6.07 (La Jolla, CA).

## Results

Participant demographics and baseline characteristics are provided in Table 1. No demographic or cell phone related variables differed significantly between groups at baseline. Parametric analysis identified one item (I sometimes miss things going on around me (i.e. conversations, class lectures, etc.) because I am doing something on my cell phone” at baseline between the comparison (2.5 ± 0.7) and the intervention (2.8 ± 0.8, *t*(210) = 2.85, *p* ≤ .005) but no other differences (*p* > .18). The internal consistency of the 12-item mobile-phone baseline assessment (questions 1 to 12 in Appendix A) was 0.78.

**Table 1.** Demographic and baseline characteristics of participants. The number of observations is in parentheses. ^A^Percent agree or strongly agree. Some items are truncated (see Appendix for full item wording).

|  | <u>Comparison</u> | <u>Intervention</u> | <u>p value</u> |
| --- | --- | --- | --- |
| Age (Mean ±SD) | 19.9 ±1.6 (59) | 20.0 ± 1.4 (155) | .53 |
| Female (%) | 62.7 (59) | 68.4% (155) | .43 |
| Smartphone ownership | 80.7% (57) | 79.9% (154) | .89 |
| I bring my cell phone everywhere I go.<sup>A</sup> | 98.3% (59) | 91.6% (155) | .08 |
| If I forgot my cell phone, I feel stressed/anxious.<sup>A</sup> | 58.6% (58) | 56.2% (153) | .75 |
| If my cell phone dies, I feel anxious.<sup>A</sup> | 56.9% (58) | 52.6% (154) | .58 |
| I check it in a setting where use is discouraged.<sup>A</sup> | 57.6% (59) | 47.7% (155) | .20 |
| My phone is one of my most important possessions.<sup>A</sup> | 53.4% (58) | 50.6% (154) | .72 |
| I miss things going on around me because of phone.<sup>A</sup> | 50.8% (59) | 36.2% (152) | .05 |
| At times I talk on the phone while driving.<sup>A</sup> | 71.2% (59) | 76.1% (155) | .46 |
| At times I text while driving.<sup>A</sup> | 43.9% (57) | 40.5% (153) | .66 |
| I sleep with my cell phone within reach.<sup>A</sup> | 88.1% (59) | 84.4% (154) | .49 |
| I view my cell phone as a necessary expense.<sup>A</sup> | 63.8 (58) | 68.8 (154) | .49 |
| I usually keep my cell phone on my person.<sup>A</sup> | 86.2% (58) | 88.9% (153) | .59 |
| I feel socially isolated without my phone.<sup>A</sup> | 32.2 % (59) | 31.4% (153) | .91 |
| I believe there may be negative social side effects.<sup>A</sup> | 81.4% (59) | 86.4% (154) | .36 |

Figure 1 depicts the mean ratings at the one-week post-test. The intervention scored significantly lower (1 = strongly agree to 5 = strongly disagree) on three of the five positively worded items (*d* = .32, .44, and .35). The fourth item, “I plan to keep my cell phone away from my body more often”, approached common statistical thresholds (*t*(213) = 1.93, *p* = .054). The two reverse worded items showed the opposite pattern with significantly higher ratings (*d* = .40 and .47). Non-parametric analyses showed the same general pattern with significant differences favoring the intervention on four items (Supplementary Figure 1).

**Figure 1.**
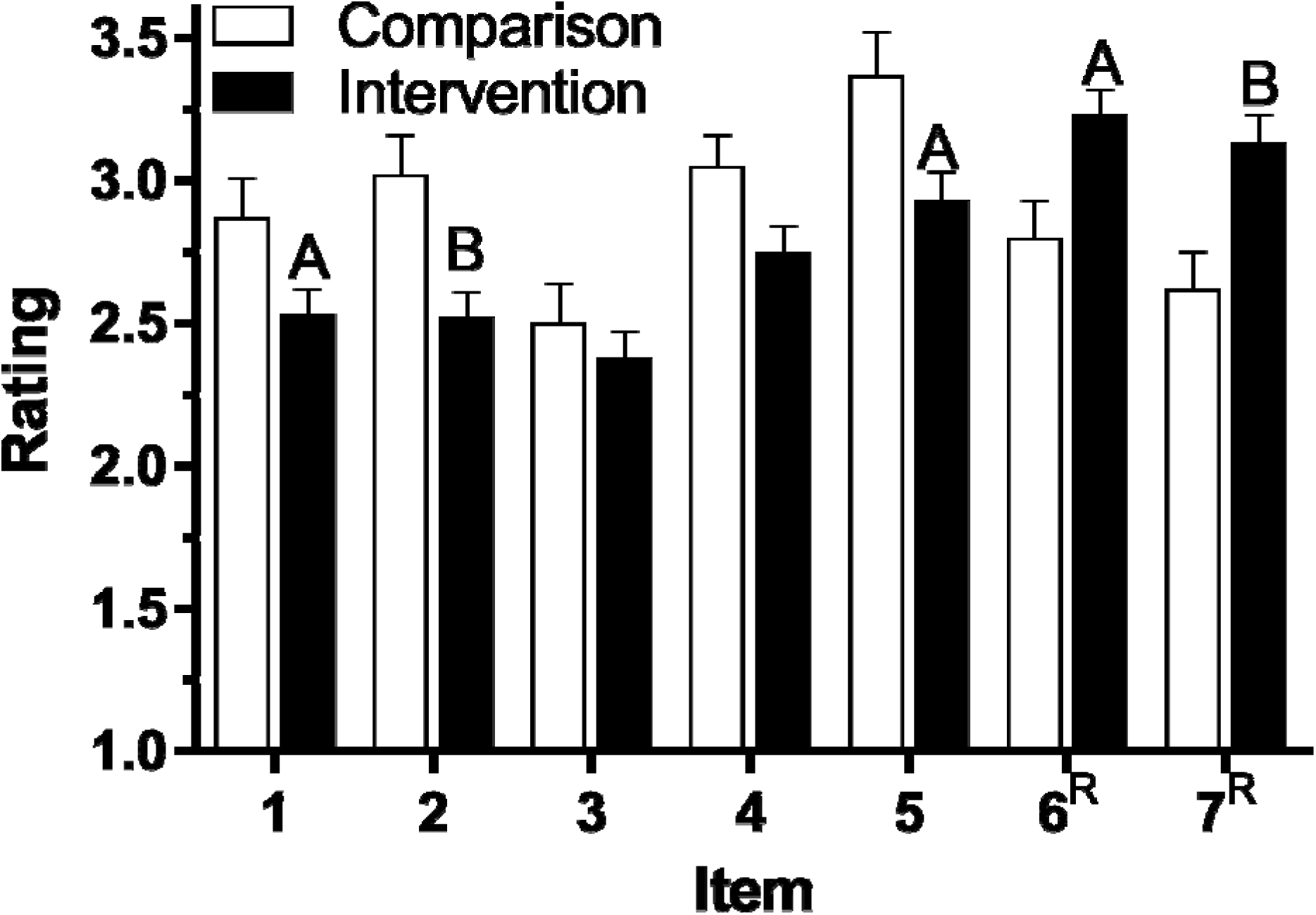
Rating on cell phone behaviors (±SEM) in emerging adults at the post-test in the Control (N = 60) and Intervention (N = 155) groups. Item 1: I am concerned that my cell phone use has negative effects on my health, 2: I am concerned that my cell phone use has negative effects on the environment, 3: I should change some of my behaviors when it comes to my cell phone use, 4: I plan to keep my cell phone away from my body more often, 5: I plan to use a hands free device for calls when I can, 6: I am *not* concerned at all about my cell phone use and behaviors, 7: I am *not* going to change the way I use my cell phone at all. ^R^reverse worded item, ^A^*p* < .05; ^B^*p* < .005 versus Comparison.

The internal consistency of the 7-item post-intervention measure was 0.82. Therefore, a total index of mobile phone behaviors was created. The Intervention scored lower than the Comparison (Figure 2, *t*(210) = 3.16, *p* < .005, *d* = .50). The effect size was larger in females (*d* = .57) than males (*d* = .38) but similar in smart-phone (*d* = .54) and non-smart-phone (*d* = .47) users.

**Figure 2.**
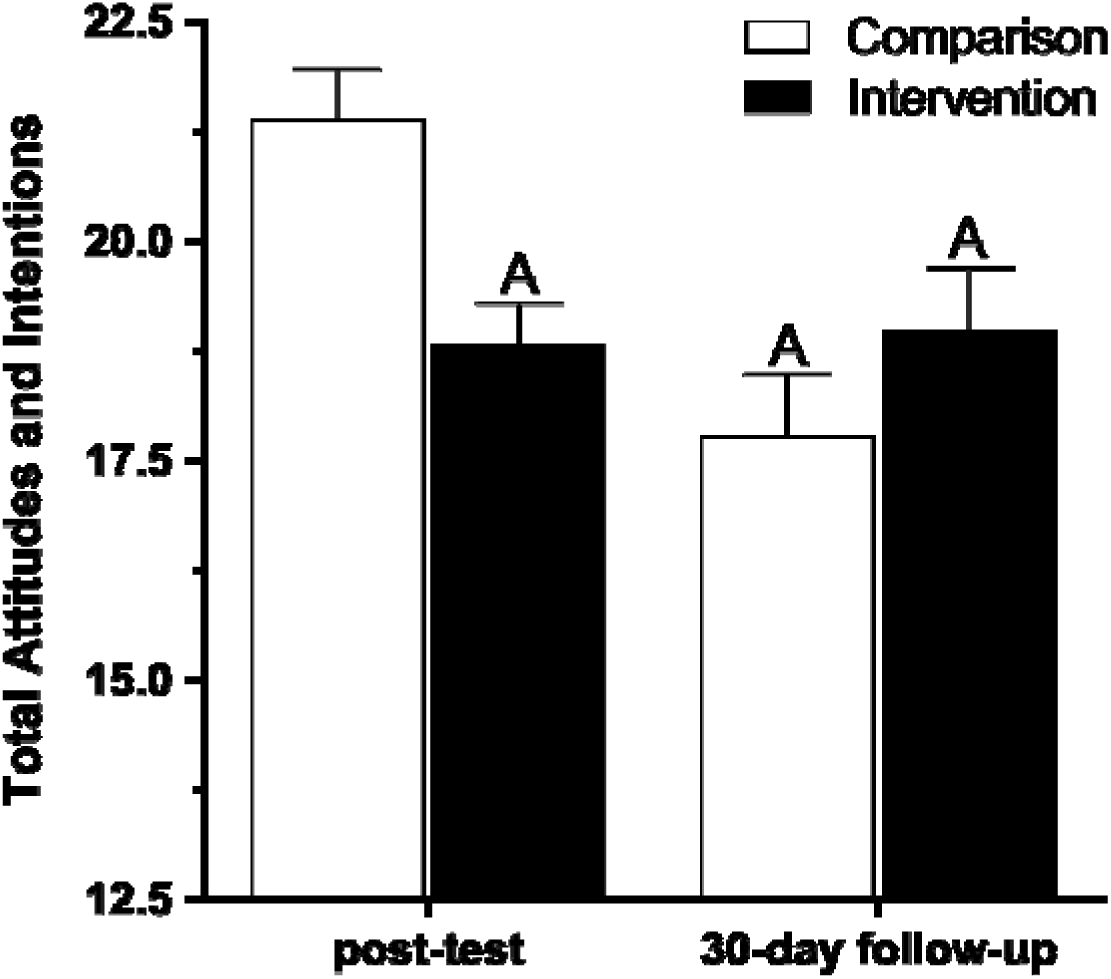
Rating on cell phone behaviors (±SEM) in emerging adults at the post-intervention (N = 212) and 30-days follow-up (N = 84) post-test. ^A^*p* ≤ .01 versus Comparison at post-test.

At one-month after the baseline, a subset of participants (33.3% of the original control and 39.4% of the intervention) were reassessed for the second Post-Test. The completers were 1.5 years younger than non-completers (Completers = 19.2 ± 1.5, Non-completers = 20.6 ± 1.3, t(213) = 7.41, *p* < .0005). More completers (79.5%) than non-completers (59.2%, χ^2^(1) = 8.85, *p* < .005) viewed their cell phone as a necessary expense but groups were otherwise indistinguishable (Supplemental Table 1). There were no differences between the intervention and comparison on individual items (*p* > .09) or the total score (*p* = .32). However, as shown in Figure 2, the Intervention at one-month was still lower than the Comparison at one-week (*t*(122) = 2.62, *p* ≤ .01). Further, the Comparison at one-month was significantly decreased relative to the Comparison at one-week (*t*(82) = 3.61, *p* ≤ .001). The test-retest correlation between post-tests at one-week and one-month was limited for all participants (*r*(84) = -.04) and within each group separately (Comparison: *r*(22) = -.14; Intervention: *r*(60) = .01). The majority of the intervention condition agreed that they learned something new from the fact-sheet at the first (87.1%) and second (65.6%) Post-Tests.

## Discussion

This report makes two contributions to the public health and behavioral addiction fields. First, this controlled study demonstrates that mobile phone use attitudes and behaviors are fluid, and perhaps, malleable in young-adults. Second, this investigation provide some preliminary psychometric support for the development of a relatively abbreviated instrument to quantify cell phone behaviors. The prevalence and use of information technology by millennials in the form of mobile devices is well established [3, 9, 21]. There is a mounting body of evidence to suggest that exposure to electromagnetic radiation emitting devices has detectable, and possibly deleterious, effects on human health and well-being [10, 12, 13, 15, 21, 47, 48, 51, 52] although see [16, 17]. Currently, the best established public-health risk associated with mobile phones may be distracted driving [25]. Young-adults, age ≤ 29 were over-represented fatal crashes involving cell phone use while driving [18]. Sleep is a fundamental neurobiological process but texting is part of a profile of poor sleep hygiene by many adolescents and young-adults [21, 50, 53]. Light-emitting electronic devices can also reduce melatonin [54]. Many people, especially those from the millennial generation, find it hard to comport their lives without continuous wireless access.

To date, there is very little available in the popular culture to alert users of any potential risks. Notably, an early association between cell phone use and brain cancer [15] did generate some media attention but this association was not substantiated in subsequent research [55]. There are only a few public policies curtailing or prohibiting mobile device use, which occur in very specific environments (e.g. texting while driving; cell phone use on an airplane or movie theater or during an educational lecture; near a gasoline pump). Risks are commonly associated only with special circumstances, in which adherence and enforcement of, may be culturally subjective. This may send the message that all other use is safe. The present findings that cell phone attitudes and behaviors can be modified, at least transiently, may be of particular interest to mental health providers with patients, or their family members, interested in reducing their mobile phone use [27]. Further, the development of cell phone interventions may be useful for individuals caught repeatedly violating laws prohibiting texting and driving.

While the majority of time spent on mobile devises is for social or recreational reasons among millennials, they also use their mobile devices as tools for all manner of information and navigation throughout their day (e.g. social media, email, clocks/calendars, GPS, audio and video entertainment). The feasibility of curtailing that multi-faceted usage has not yet been well established. This investigation identified a small-moderate effect size with a brief-intervention at a short (one-week) interval.

Although not the primary objective, this study also adds to other findings to develop other abbreviated measures of problematic cell phone use [56–59]. A Smartphone Addiction Inventory has been developed for cell phone users that can read Mandarin which showed excellent internal consistency and very good two-week test-retest reliability in a primarily male young-adult sample [60]. The internal consistency of both the baseline and Post-Test instrument used in this effort was sufficient. However, this was not the case for the test-retest reliability. However, as only a minority of participants were available for the one-month Post-Test, any inferences regarding the data at this interval (correlations or means) should be made with great caution.

Some additional limitations and future directions are also noteworthy. First, although the participants in this controlled study provided evidence that they changed their cell phone use behaviors, and this post-intervention pattern appeared to be persistent (relative to the comparison group at one-week), the endpoints measured were based on self-report. Future efforts to reduce phone use could also incorporate more objective measures like phone bills or an application that records mobile phone use [61]. Second, the participants were young-adults and disproportionately female. As high-school and college-age students have the highest mobile phone usage [21], and some data indicates that females have more problematic cell phone use patterns [50, 53], we view the sample characteristics as a strength of this report. However, additional longitudinal studies will be necessary to further characterize the persistence of mobile phone attitudinal and behavior changes following interventions. This investigation was completed with a normative sample of young-adults. More intense, and repeated, interventions may be needed for individuals with more excessive, and problematic, mobile-phone attitudes and behaviors [27].

In conclusion, this investigation provides evidence that cell phone use can be modified, at least temporarily, in young-adults.

## Data Availability

Data is included as a supplemental material.

## Acknowledgments

The efforts of the peer educators are gratefully acknowledged.

---

**Supplemental Table 1.** Demographic and baseline characteristics of participants that did (completers), and did not (non-completers), participate in the one-month Post-Test. The number of observations is in parentheses. Some items are truncated (see Appendix for full item wording). ^A^Percent “agree” or “strongly agree”.

|  | <u>Completers</u> | <u>Non-completers</u> | <u>p value</u> |
| --- | --- | --- | --- |
| Age: Mean ±SD | 19.2 ± 1.5 (88) | 20.6 ± 1.3 (127) | < .0005 |
| Female | 63.6% (88) | 69.3% (127) | .47 |
| Smartphone ownership (%) | 81.8% (88) | 78.2 (124) | .64 |
| I bring my cell phone everywhere I go.<sup>A</sup> | 94.3% (88) | 92.9% (127) | .90 |
| If I forgot my cell phone, I feel stressed or anxious.<sup>A</sup> | 58.6% (87) | 56.0% (125) | .81 |
| If my cell phone dies, I feel stressed or anxious.<sup>A</sup> | 56.8% (88) | 52.0% (125) | .58 |
| I check it in a setting where cell phone use is discouraged.<sup>A</sup> | 51.1% (88) | 49.6% (127) | .94 |
| My cell phone is one of my most important possessions.<sup>A</sup> | 52.3% (88) | 50.4% (125) | .90 |
| I sometimes miss things because of my phone.<sup>A</sup> | 44.7% (85) | 37.0% (127) | .33 |
| I talk on the phone while driving.<sup>A</sup> | 77.3% (88) | 72.4% (127) | .52 |
| I text on the phone while driving.<sup>A</sup> | 38.4% (86) | 43.2% (125) | .58 |
| I sleep with my cell phone within reach.<sup>A</sup> | 82.8% (87) | 87.4% (127) | .45 |
| I view my cell phone as a necessary expense.<sup>A</sup> | 79.5% (88) | 59.2% (125) | .003 |
| I usually keep my cell phone on my person.<sup>A</sup> | 83.9% (87) | 91.2% (125) | .16 |
| I feel socially isolated when I don't have my phone.<sup>A</sup> | 31.4% (86) | 31.5% (127) | 1.00 |
| There may be negative social side effects to cell phones.<sup>A</sup> | 86.2% (87) | 84.3% (127) | .84 |

**S1 Appendix.**
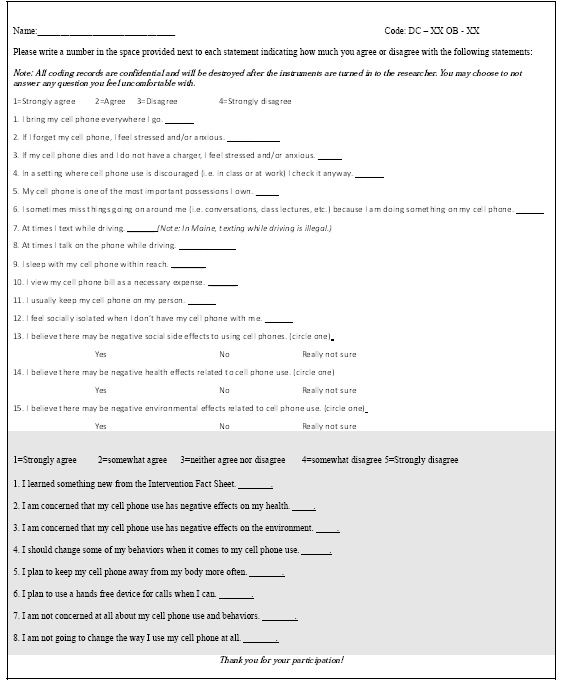
Pre and Post Intervention Survey

**S Figure 1.**
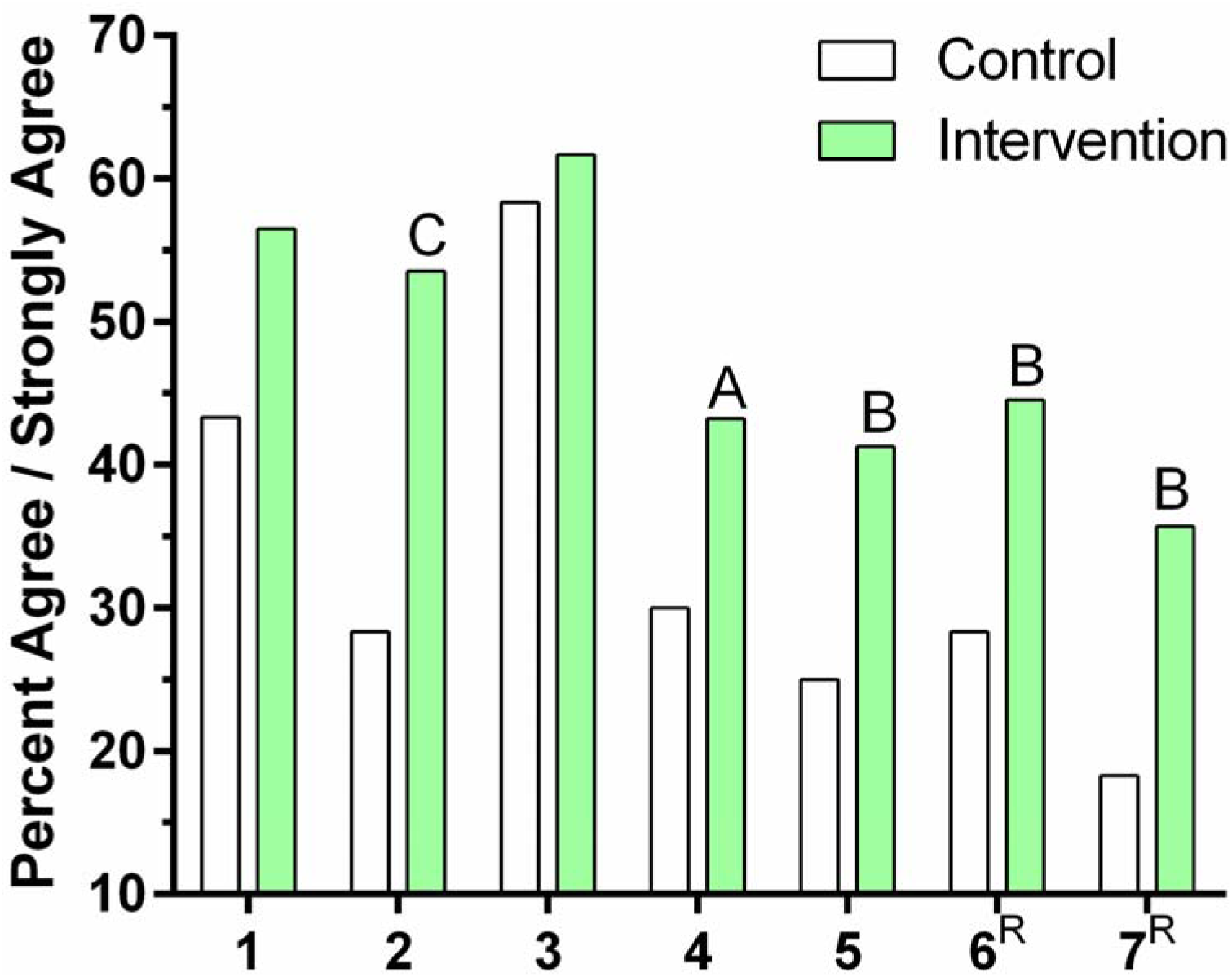
Percent agreement on cell phone behaviors at the one-week post-test in the Control (N = 60) and Intervention (N = 155) groups. Item 1: I am concerned that my cell phone use has negative effects on my health. Item 2: I am concerned that my cell phone use has negative effects on the environment. Item 3: I should change some of my behaviors when it comes to my cell phone use. Item 4: I plan to keep my cell phone away from my body more often. Item 5: I plan to use a hands free device for calls when I can. Item 6: I am *not* concerned at all about my cell phone use and behaviors. Item 7: I am *not* going to change the way I use my cell phone at all. Items were rated 1= strongly agree, 2 = agree, 3 = neutral, 4 = disagree, and 5 = strongly disagree except items 6 and 7 which were ^R^reverse coded. ^A^*p* < .10; ^B^*p* < .05; ^C^*p* < .005.

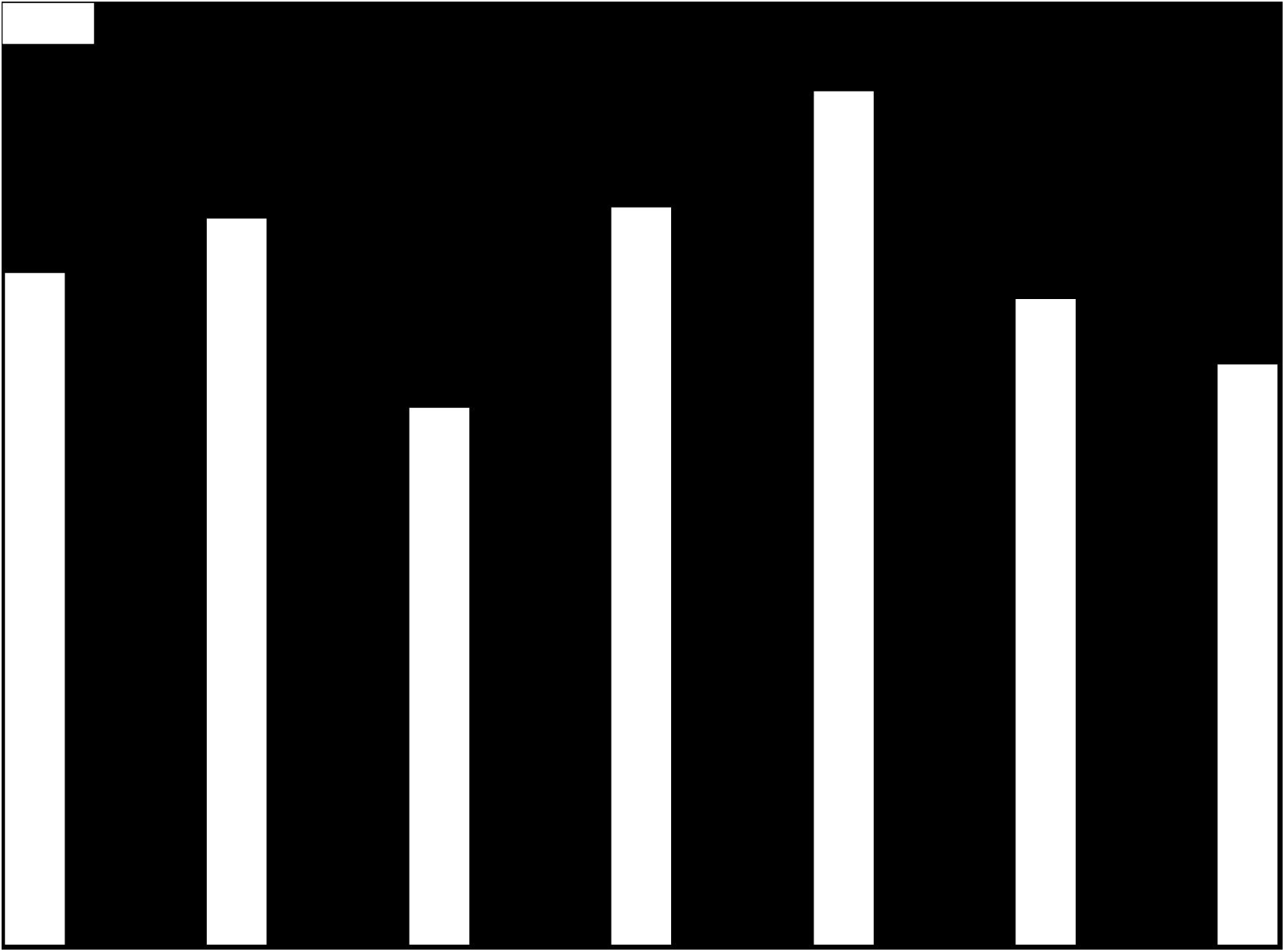

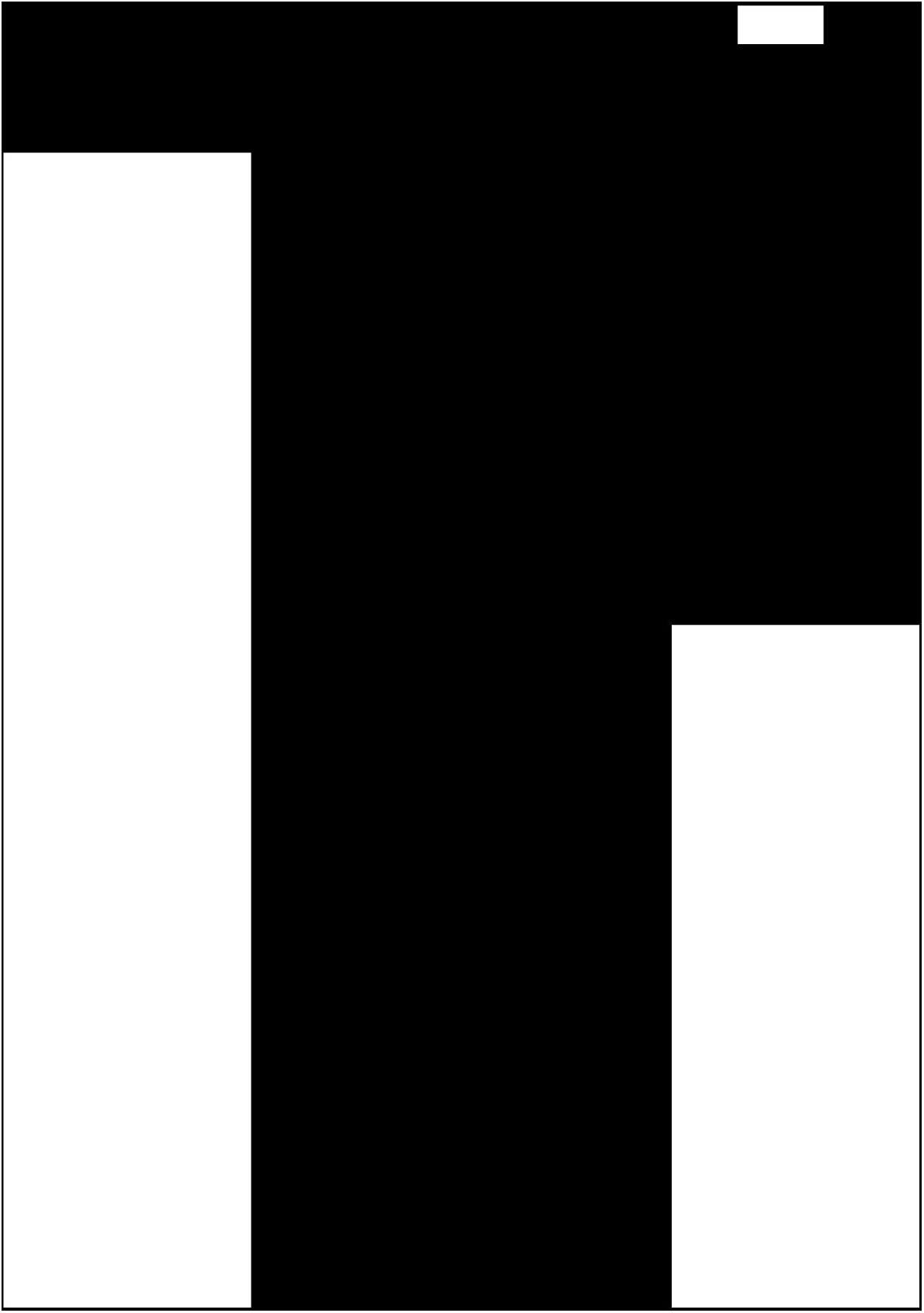

